# Smartphone-based measurement of cognition and physical function in older emergency department patients: A Feasibility Study

**DOI:** 10.1101/2024.10.24.24316067

**Authors:** Andrew Leroux, Roland C. Merchant, Samantha Roberts, Brianne M. Bettcher, Hillary D. Lum, Linda Resnik, Sarah D. Berry, Meredith Mealer, Vince Mor, Elizabeth Goldberg

**Affiliations:** Department of Biostatistics and Informatics, Colorado School of Public Health, Aurora, CO, USA; Department of Emergency Medicine, Icahn School of Medicine at Mount Sinai, New York City, NY, USA; Department of Neurology, University of Colorado Anschutz Medical Campus, Aurora, CO, USA; Division of Geriatric Medicine, Department of Medicine, University of Colorado Anschutz Medical Campus, Aurora, CO, USA; Center of Gerontology and Healthcare Research, Brown University School of Public Health, Providence, RI, USA; Hinda and Arthur Marcus Institute for Aging Research, Hebrew SeniorLife, Boston, MA, USA; Department of Medicine, Beth Israel Deaconess Medical Center & Harvard Medical School, Boston, MA, USA; Department of Emergency Medicine, University of Colorado Anschutz Medical Campus, Aurora, CO, USA

**Author notes:** **CORRESPONDING AUTHOR:** Elizabeth Goldberg, MD, ScM, University of Colorado, Department of Emergency Medicine.

**Keywords:** Falls, digital measures, emergency department, geriatrics, wearable devices, clinical trial, smartphone, Apple Watch, functional assessment, feasibility study, cognitive assessment

## Abstract

**Background:** Wearables and smartphones are increasingly deployed in digital assessments of cognitive and physical function in studies of older adults. However, their use in the emergency department (ED) for patients presenting with falls has been limited. In GAPcare II, a randomized-controlled trial of an ED-based fall prevention intervention, we combined standard quantitative measures to screen for cognitive impairment and physical function limitations, with digital measures of cognition and physical function using smartphones and smartwatches. Our objective was to assess the feasibility of deploying digital assessments in the ED for older patients with a recent fall.

**Methods:** Between August 2021 and January 2024, community-dwelling older (≥ 65 years old) ED patients presenting for a fall were screened for cognitive impairment (Six Item Screener) and physical function limitations (use of mobility equipment, modified Barthel Index). Apple ResearchKit digital assessments were administered using smartphones and smartwatches to assess cognition (Stroop, Trail-Making tests, reaction time) and physical function (gait and balance, timed walk test). Wearable devices were applied to the patient’s wrist for passive movement measurement. Patients were instructed and supervised by trained research staff. We assessed feasibility by determining how many patients attempted and completed each digital task, along with reasons for non-attempt and non-completion. We also assessed the association between test completion and patient characteristics in univariate and multivariable regression models.

**Results:** Among 197 patients, the average age was 78.2 years (standard deviation = 7.6), and 68% were women. Twelve percent had possible cognitive impairment, and 70% had some functional dependence. Eighty-two percent attempted at least one digital task. Leading reasons for non-attempt included concerns surrounding safety and pain or discharge from the ED before the attempt, specifically for the physical tasks. Completion rates among those who attempted were moderately high (68-87%) for cognitive tests and did not vary by age, other demographic variables, or health behaviors (e.g., tobacco, alcohol use), but did vary by possible cognitive impairment (p<0.01, all cognitive tests) and physical function (Barthel index, p<0.01, reaction time test only). Reasons for non-completion of cognitive tests included injury (15%), task was “too hard” (7%), and technology issues (7%). Completion rates for physical function tasks were substantially lower (18-20%) and did not vary by demographic characteristics but did vary with standard measures of physical function (Barthel Index and use of mobility equipment, p<0.01 and p<0.05, respectively). Low completion rates for physical function tests were mainly due to safety concerns, pain, and injury.

**Conclusions:** Digital assessment of cognitive function using publicly available smartphone-based tests is feasible in studies of older adults presenting to the ED for falls. However, patients may be reluctant to engage in physical function tests requiring mobilization immediately after an injury. Future research will investigate whether such data are predictive of clinically relevant outcomes (e.g., time to injury recovery, ED return visits) and can inform ED care (e.g., referrals to physical therapy, skilled nursing facility placement).

## Background

Falls are the leading cause of injury-related mortality in older adults, ≥65 years old.^1^ Older adults presenting to the emergency department (ED) for a fall-related injury are at substantial risk of short-term subsequent falls, institutionalization, and mortality.^2, 3^ A fall-related ED visit provides an opportune time to intervene during the ED visit to reduce modifiable risk factors for subsequent falls, given the risk for a rapid health decline post-fall, and because of the tendency of older adults to underreport falls^4, 5^ to primary care clinicians in follow-up after the ED visit. Assessment of fall risk factors, such as cognitive and physical function during the ED visit, is critical to personalizing interventions. Unfortunately, few older adults visiting the ED after a fall currently benefit from these assessments because they often are reliant on the availability of trained clinical staff (e.g., physical therapists and occupational therapists) to perform them.

Mobile health (mHealth) measures of cognition and physical function are increasingly available and may enable bedside clinical assessment when highly trained clinical staff are unavailable. Although these digital measures do not replace traditional neuropsychological and physical function testing, they can complement current assessment methods. In addition to their use in clinical settings, digital assessments might be particularly appealing in longitudinal studies during which investigators study intraindividual cognitive and functional variability,^6^ and in studies conducted remotely or in settings where assessors are unavailable. Poor performance on cognitive measures that assess executive function, such as reaction time, and on physical function measures, such as gait speed, have been associated with future fall risk^6-9^ and digital versions of these measures have been made publicly available for use on smartphones. Despite the rapid developments in mHealth measures of cognition and physical function, the feasibility of deploying them in the ED, particularly among older patients after a fall, remains unknown.

The primary objective of this study was to evaluate the feasibility of using digital technology, specifically smartphones, in conjunction with wrist-worn wearable devices to assess cognitive and physical function in the ED among older patients presenting for an evaluation after a fall. We hypothesized that conducting both cognitive and physical tests via digital technologies was feasible in this setting. The second, exploratory objective of this study was to measure the association of digital assessment outcomes (Stroop effect, accuracy of Stroop test items, Trail-Making test time, number of errors made during the Trail-Making test) with age, other demographic characteristics, validated measures of cognition (six-item screener) and physical function (use of mobility equipment, Barthel Index), prior falls, and injury recovery time post-ED visit. We hypothesized that older age, possible cognitive impairment, physical limitations, prior falls, and prolonged injury recovery times would be associated with lower scores on digital cognitive measures.

## Materials and Methods

### Study Design and Setting

Data was collected from patients enrolled in the GAPcare II randomized-controlled trial (The Geriatric Acute & Post-acute Care Coordination Program for Fall Prevention in the Emergency Department). From August 2021 to January 2024, GAPcare II recruited 197 patients presenting to the ED for a fall at two hospitals in Rhode Island, The Miriam Hospital (n=47) and Rhode Island Hospital (n=71), as well as at the University of Colorado Hospital in Colorado (n=79).

The GAPcare II protocol provides more details on inclusion and exclusion criteria and procedures.^10^ Briefly, patients were study eligible if they were community-dwelling (not institutionalized), were ≥65 years old, and presented for a fall. Patients performed written informed consent. Patients lacking the capacity to complete informed consent were offered proxy consent. Upon successful recruitment, patients were asked to complete several surveys, and those randomly assigned to the intervention arm received an in-ED pharmacy and physical therapy consultation.

Following these assessments, the research staff asked patients to complete the digital assessments. Staff members introduced each task and were available to help guide the patient during the duration of the tasks. First, the research staff member provided the patient with an Apple Watch (series 4) and an iPhone (version 12). The Apple Watch was always applied to the non-dominant wrist. Next, research staff instructed the patient to perform five digital assessments: three that assess cognition, followed by two that assess physical function. All digital assessments were Active Tasks deployed through ResearchKit, an open-source framework for health applications. All patients enrolled in the GAPcare II study were eligible for inclusion in the current analysis.

### Cognitive Tests

Cognitive tests were deployed on iPhones running MacOS version 15-17 (2024, Apple Inc.) were connected to Apple Watch devices. Figure 1 illustrates each of these tests from the patient’s perspective. Auditory, tactile, and visual cues accompanied tests. Each test had written instructions visible on the screen. A digital voice also relaid instructions, and the user was prompted by vibration from the device when an action was needed. We made no modifications to the digital versions of the test provided through Apple’s ResearchKit, although the digital versions differ from the traditional in-person versions in several ways, as described below. These digital versions have not been validated against “gold standard” tests, which are recommended to be administered in written format by a neuropsychologist or other trained personnel.

**Figure 1.**
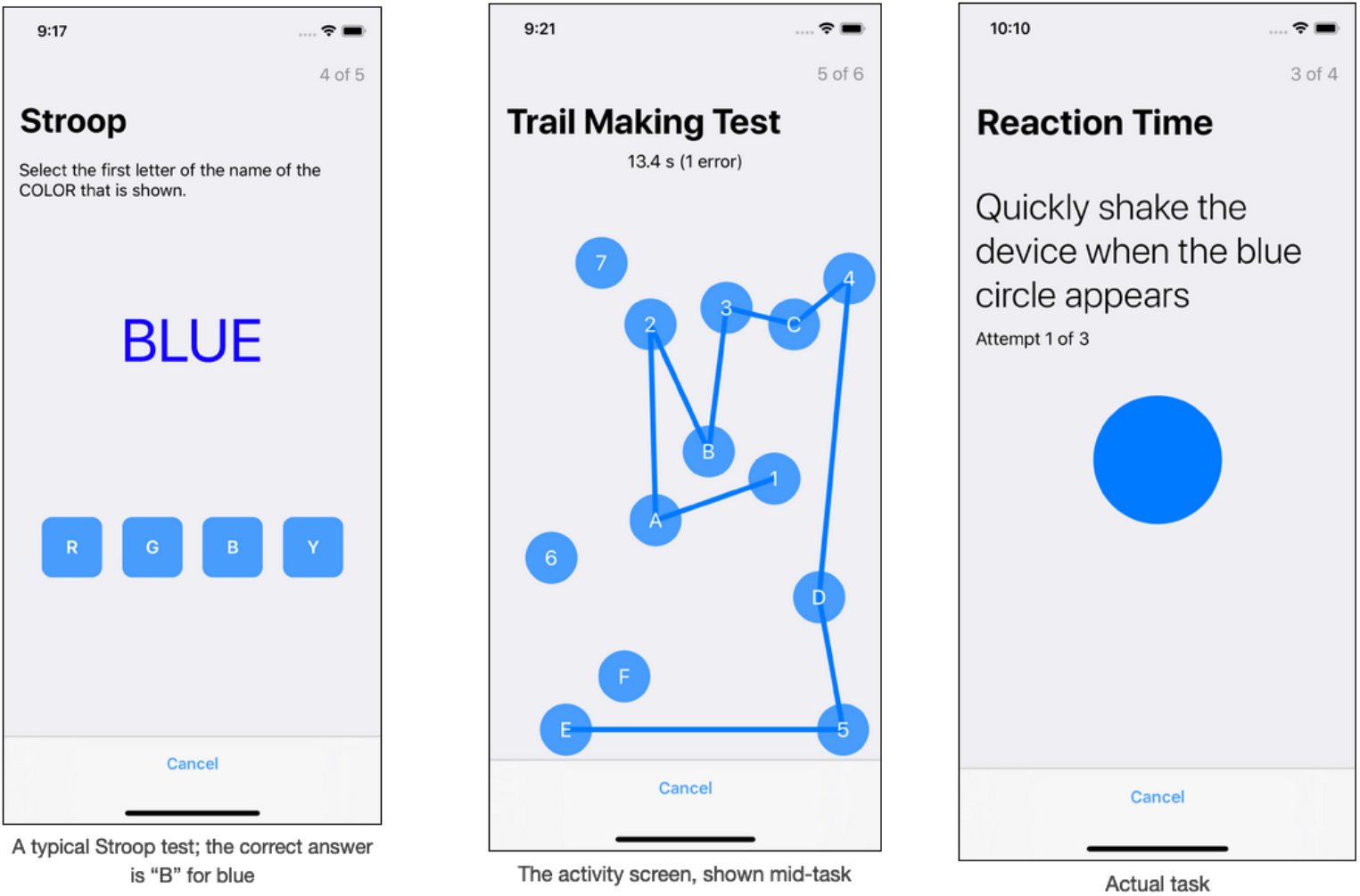
Illustration of the Cognitive Tests (Stroop test, Trail-Making test and reaction time test, top left, middle, and right panels, respectively) and instructions for the physical function tests (gait and balance and timed walk tests, bottom left and right panels, respectively).

#### Stroop Test

The Stroop test deployed in our study presented patients with a series of words, one of: blue, yellow, red, or green. The font color of the word either matched the word (e.g., “blue” shown in blue font) or differed (e.g., “blue” shown in yellow font). Patients were asked to select the first letter of the name of the color that was shown (see left panel of Figure 1). Patients were presented with 15 different word/color combinations, with each pair randomly selected. Patients were shown different numbers of congruent pairs, ranging from 3 to 11. This Stroop test differed from standard variants in several ways. The classical variants of the Stroop test ask patients to read aloud the word shown, and patients are given an equal number of congruent/incongruent pairs. To analyze the Stroop test, we considered the average time to respond for congruent and incongruent color/word pairs, differences in patients’ average response time for congruent versus incongruent pairs (the Stroop effect), and the percent of correct responses by congruent vs. incongruent pairs.

#### Trail-Making test

The Trail-Making test given to patients required them to connect dots on the screen in alternating numeric and alphabetical order (1-A-2-B-…-6-F-7), a total of 13 connections. If patients selected incorrectly, they were allowed to continue the test from their last correct guess. This version of the Trail-Making test differed from traditional versions in two key ways. First, traditional Trail-Making tests first ask patients to connect consecutive numbers without any letters present (e.g., 1-2-…-25). This test is followed by one that requires patients to sequence alternating numbers and letters. Additionally, the letter-number portion of a classical Trail-Making test involves numbers from 1-13 and letters A-L, resulting in a test with more connections (25 total). To analyze the Trail-Making test, we considered the total time to complete the test and the total number of incorrect guesses made.

#### Reaction Time Test

The reaction time test asks patients to shake the phone (in any direction) as soon as a large blue circle appears on the screen. The time until the blue dot appears is varied. As soon as the device registers movement at sufficiently high acceleration, the task is complete. Patients are asked to repeat this three times, with time to completion for each reaction stored.

### Physical Function Tests

Physical function tests were deployed using the same devices as were used for the cognitive tests.

#### Gait and Balance Test

For the gait and balance test, patients’ gait and balance are assessed while standing still and walking. Users are prompted by their iPhone to walk 20 steps in a straight line, turn around, walk back to their starting position, and then stand still for 20 seconds. Motion is assessed using the device’s accelerometer and gyroscope. For the GAPcare II study, the phone was secured to patients by a transparent fanny pack around the waist with the iPhone screen visible to the staff.

#### Timed Walk Test

Following a prompt from the phone, patients were asked to walk a fixed distance as quickly as they could safely. Once the device had determined the appropriate distance has been walked, patients were prompted to turn around and walk to their starting position. Distance and movement were assessed using the device’s gyroscope and GPS. Patients were asked to walk 109 yards in the ED hallway, accompanied by study staff. Devices were secured to patients in the same fashion as the gait and balance test. Users could indicate if they plan to use an assistive device for walking before beginning the test.

### Feasibility Outcomes

Our major objective was determining feasibility. We assessed feasibility by calculating attempt and completion rates for each of the tasks and determining reasons for non-attempts and non-completion, described as follows:

#### Attempt

The patient was available and interested in starting at least one digital assessment.

#### Reasons for non-attempt

Reasons for patient non-attempt were assessed and categorized.

#### Completion

The completion of a test (yes/no) was determined by whether a patient agreed to participate and whether cognitive or functional assessment data were successfully collected. We also recorded whether any digital measure was attempted, the reasons for non-attempt, and, if attempted, the reasons for non-completion or early stoppage. The completion of each test was calculated separately. Research staff recorded technical problems (e.g., device and connectivity issues) and challenges they observed.

#### Reasons for non-completion

Reasons for non-completion were assessed and grouped into five categories (technology problem, patient refusal prior to attempt, patient aborted after trying, patient injury prevented completion, patient left ED prior to completion). Within broad categories, the reason for non-completion was broken down into several sub-categories.

#### Clinical characteristics

We considered the association of each outcome with demographic characteristics and clinically relevant factors. Specifically, we examined associations of the outcome measures with age, race/ethnicity, body mass index (BMI), gender (man/woman), education (some grade school, high school equivalent, some college, college graduate), cigarette smoking (yes/no), recent alcohol consumption prior to fall (yes/no), use of an assistive device for movement (yes/no assistive device use), functional impairment/independence (Barthel index < 20,=20), cognitive impairment (six-item screener score <4,4-6), self-reported history of a fall within the past three months (yes/no), and time to return to normal function following the ED visit (return to normal within 3 days, return to normal longer than 3-30 days, still not normally functioning at 30 days). Time to return to normal function was assessed at the 30-day follow-up survey, conducted over the phone or in person at the patients’ homes.

### Statistical Analysis

We categorized reasons for nonattempt and noncompletion and the frequency of their occurrence.

Distributions of outcomes were evaluated using proportions (binary outcomes, completion rates) and summary statistics (continuous outcomes, cognitive and function test results). Conditional associations were estimated using generalized additive models with non-linear associations between each outcome and age estimated using rank ten penalized cubic regression splines.^11, 12^ Gaussian, logistic, and Poisson regression were used for continuous, binary, and count data, respectively. Wald-type confidence intervals for regression coefficients were constructed. For regressions with completion as an outcome, the area under the receiver operating characteristic curve statistics was calculated along with 95% confidence intervals.^13^ Multivariable regression models included a common set of variables across models except for the Stroop effect and Trail-Making test time. These outcomes additionally adjusted for the accuracy of incongruent and congruent pairs, as well as the number of congruent pairs (Stroop effect), and the number of errors made in the Trail-Making test (Trail-Making test time). Analyses were exploratory, and as such, no multiple comparison adjustments were made for statistical tests of significance.

## Results

We recruited 197 patients; nearly half were 74 to 85 years old (n=94, 48%), and the majority were women (Table 1). Most patients were non-Hispanic/Latino and white. Almost all patients had minimal risk of cognitive impairment, Six-Item Screener >3, with increasing impairment prevalence in older age groups (p<0.001). The majority of patients reported some level of functional dependence and used assistive devices for mobility. Patients were generally non-smokers and did not consume alcohol preceding the fall. Approximately half of patients reported at least one fall in the past 3 months, exclusive of the ED visit fall, and the majority reported being injured in their fall that led to the ED visit. Most patients reported having more than a high school education, with the level of education varying by age (p=0.016).

**Table 1.** Demographics and clinical characteristics of patients.

| Characteristics |  | Age Group |  |  | p-value |
| --- | --- | --- | --- | --- | --- |
|  | Overall | (64,74] | (74,85] | (85,100] |  |
|  | 197 | 71 | 94 | 31 |  |
| **Gender** |  |  |  |  | 0.269 |
| Female | 134 (68.0) | 52 (73.2) | 59 (62.8) | 23 (74.2) |  |
| Male | 63 (32.0) | 19 (26.8) | 35 (37.2) | 8 (25.8) |  |
| **Ethnicity** |  |  |  |  | 0.689 |
| Not Hispanic/Latino | 179 (90.9) | 64 (90.1) | 86 (91.5) | 28 (90.3) |  |
| Hispanic/Latino | 12 ( 6.1) | 6 ( 8.5) | 4 ( 4.3) | 2 ( 6.5) |  |
| Unknown | 6 ( 3.0) | 1 ( 1.4) | 4 ( 4.3) | 1 ( 3.2) |  |
| Cognitive impairment<br>(Six Item Screener, SIS) |  |  |  |  | <0.001 |
| SIS, score < 4 (concern<br>for cognitive impairment) | 23 (12.2) | 1 ( 1.5) | 13 (14.1) | 9 (31.0) |  |
| SIS, score 4-6 (within<br>normal range) | 166 (87.8) | 66 (98.5) | 79 (85.9) | 20 (69.0) |  |
| **Barthel Index (BI)** |  |  |  |  | 0.125 |
| BI = 20 (Complete<br>Independence) | 59 (30.1) | 21 (29.6) | 33 (35.5) | 5 (16.1) |  |
| BI < 20 (Some<br>Dependence) | 137 (69.9) | 50 (70.4) | 60 (64.5) | 26 (83.9) |  |
| **Cigarette smoking** |  |  |  |  | 0.256 |
| Non-smoker | 188 (96.9) | 67 (94.4) | 90 (98.9) | 30 (96.8) |  |
| Smoker | 6 ( 3.1) | 4 ( 5.6) | 1 ( 1.1) | 1 ( 3.2) |  |
| **Education** |  |  |  |  | 0.016 |
| Less than High School | 29 (15.1) | 7 (10.0) | 18 (20.0) | 4 (12.9) |  |
| High School | 42 (21.9) | 12 (17.1) | 17 (18.9) | 13 (41.9) |  |
| More than High School | 121 (63.0) | 51 (72.9) | 55 (61.1) | 14 (45.2) |  |
| **Alcohol consumption**<br>**prior to fall** |  |  |  |  | 0.848 |
| Nondrinker | 158 (80.6) | 59 (83.1) | 74 (79.6) | 25 (80.6) |  |
| Drinker | 38 (19.4) | 12 (16.9) | 19 (20.4) | 6 (19.4) |  |
| **Injured during ED**<br>**index visit fall** |  |  |  |  | 0.900 |
| No | 21 (10.8) | 7 ( 9.9) | 11 (12.0) | 3 (10.0) |  |
| Yes | 173 (89.2) | 64 (90.1) | 81 (88.0) | 27 (90.0) |  |
| **Use Mobility**<br>**Equipment** |  |  |  |  | 0.061 |
| No | 60 (30.6) | 25 (35.2) | 31 (33.3) | 4 (12.9) |  |
| Yes | 136 (69.4) | 46 (64.8) | 62 (66.7) | 27 (87.1) |  |
| **Any falls in past 3**<br>**months** |  |  |  |  | 0.478 |
| No Falls | 93 (47.4) | 30 (42.3) | 48 (51.6) | 14 (45.2) |  |
| 1+ Falls | 103 (52.6) | 41 (57.7) | 45 (48.4) | 17 (54.8) |  |
| **Time to return to function after ED index visit fall** |  |  |  |  | 0.780 |
| No Reduction | 23 (15.1) | 10 (16.4) | 12 (17.4) | 1 (4.8) |  |
| Less than 3 days | 30 (19.7) | 12 (19.7) | 14 (20.3) | 4 (19.0) |  |
| More than 3 days, back to normal | 34 (22.4) | 12 (19.7) | 15 (21.7) | 7 (33.3) |  |
| Still not full function at 30 days | 65 (42.8) | 27 (44.3) | 28 (40.6) | 9 (42.9) |  |
| **Heart Rate** | 76.87 (16.43) | 78.95 (19.16) | 74.91 (12.41) | 80.10 (20.52) | 0.454 |

p-values calculated using t-test for continuous variables and Fisher's exact test for categorical variables.

### Feasibility: Attempt and Completion rates

Table 2 presents patient reasons for non-completion for cognitive tests (Table 2A) and physical function tests (Table 2B) by category (rows) and test (columns). Tables 2A/2B are stratified by attempters versus non-attempters. Some reasons for non-attempt and non-completion were task specific. Of the 197 patients, 77% (n=151) attempted the digital measures. Reasons for non-attempt varied by test and included the patient declined, the patient feared it was not safe or had pain, the patient was discharged from the ED before the task attempt, the patient having other illness(es) that prevented the task attempt, too tired, and the patient already disenrolled from the study (n=1). Of those who attempted, some met technology challenges such as the app crashing, the app failing to record data, and the app being unavailable for download. Ninety-nine percent of those who attempted (n=138) and did not meet technology challenges, completed at least one test. Completion of the Stroop and Trail-Making tests were high among attempters at 87% and 84%, respectively. The completion rate for the reaction time test was lower at 68%. The physical function tasks had lower completion rates, with 20% and 18% of attempters completing the gait and balance and timed walk tests, respectively.

**Table 2.**
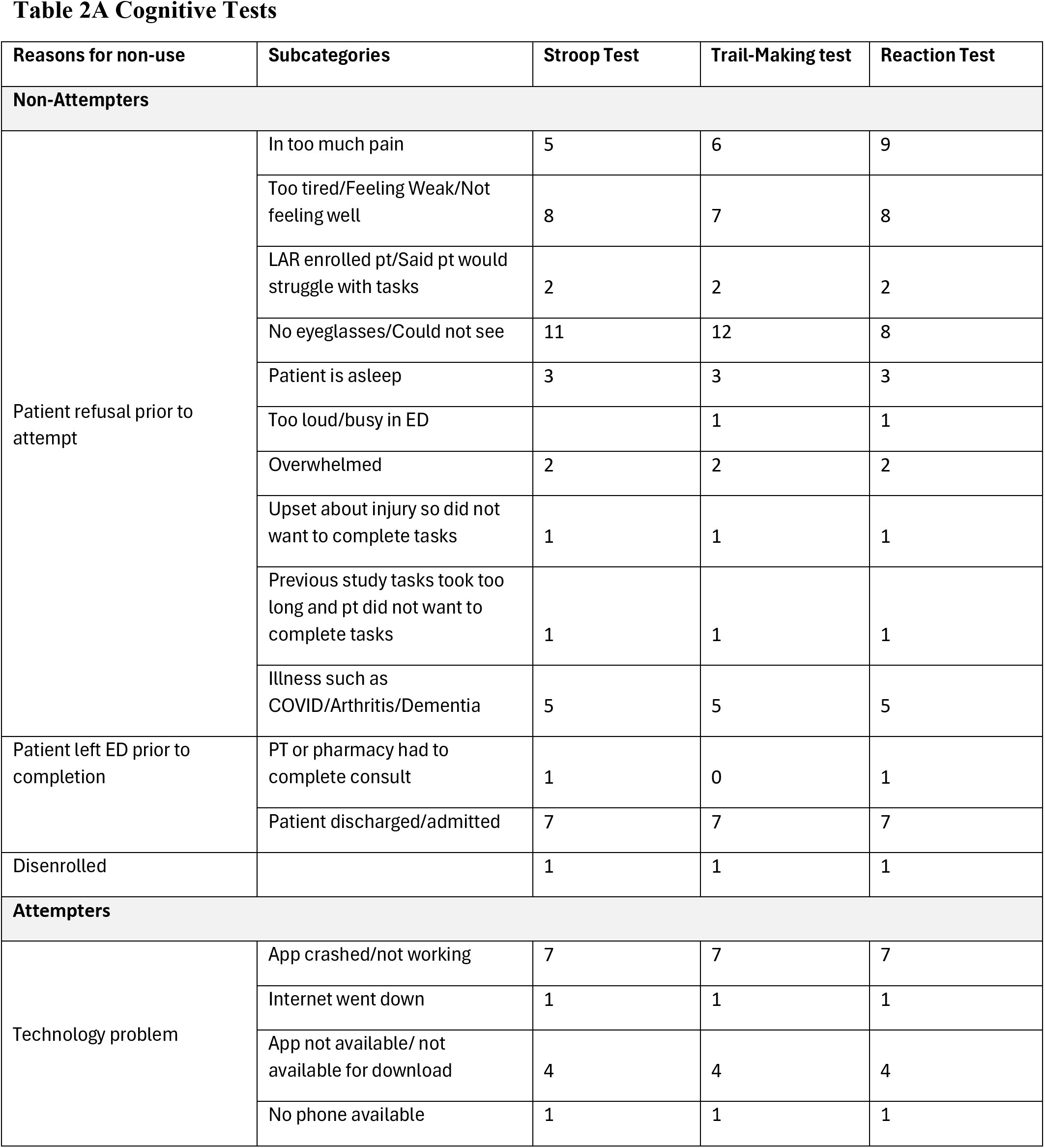
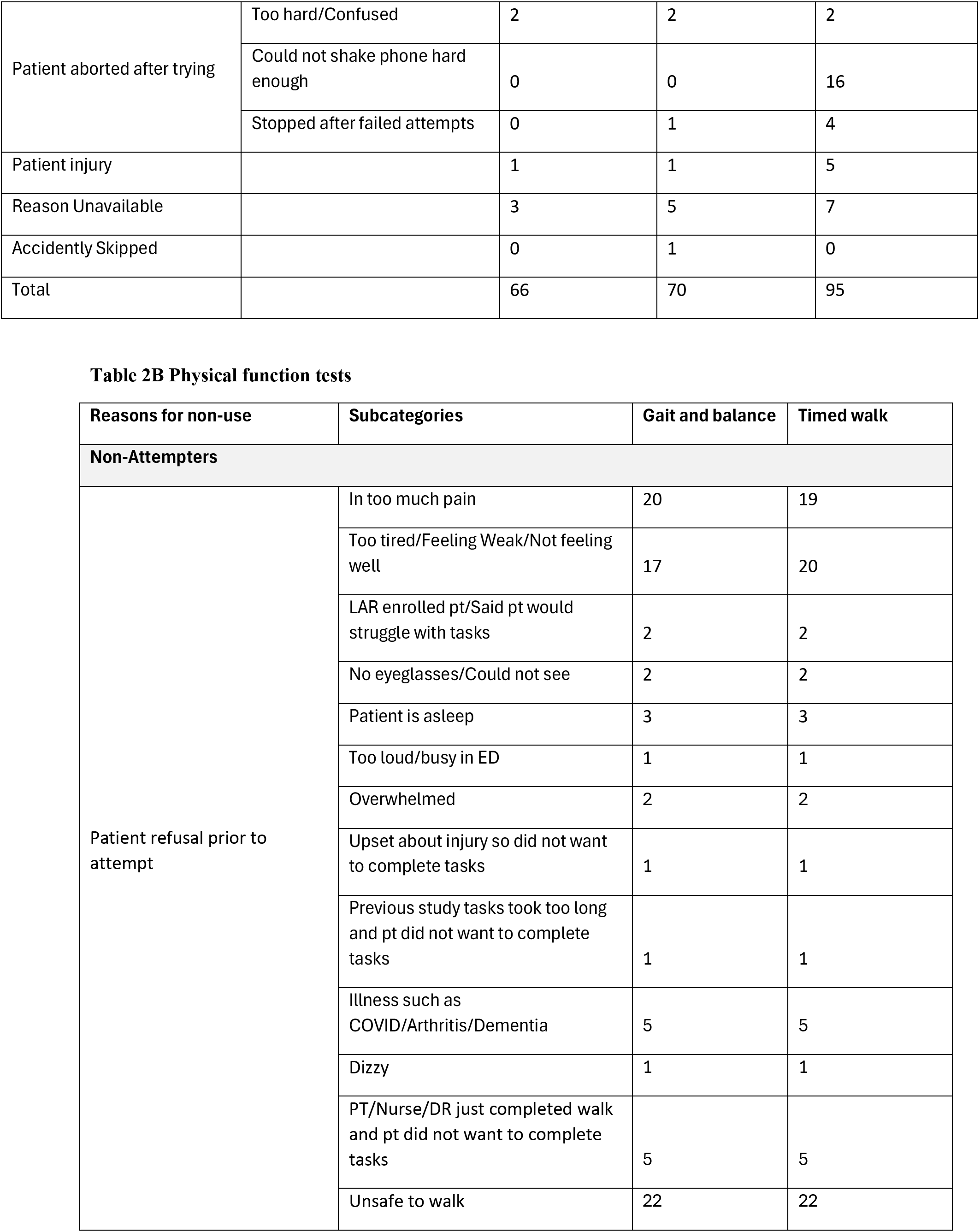
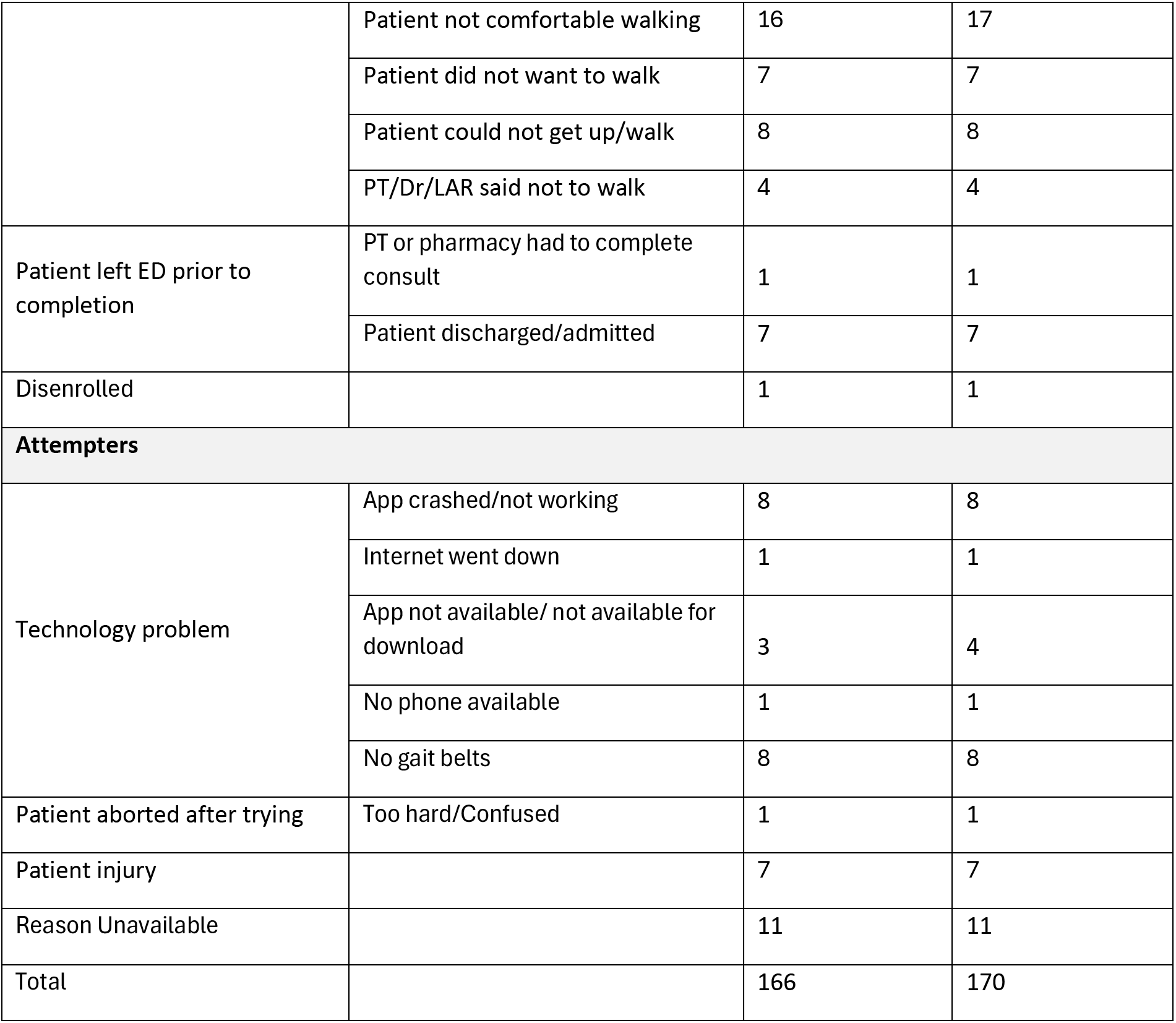
Reasons for patient non-completion of digital assessments for cognitive tests (Table 2A) and physical function tests (Table 2A)

### Reasons for non-completion

Of the 151 patients who attempted the measures, the most common reasons for not completing the physical function tasks were patients’ reports of not feeling well enough to perform the tasks or being worried about their ability to do so safely, particularly for the physical function tasks. Physical function tests required the patient to get out of the stretcher and walk in the ED hallway accompanied by staff. Other major reasons for non-completion included injury or the test being too hard. Other patients cited desiring to leave the ED, being in too much pain, being too tired, or lacking eyeglasses. Generally, if patients aborted the reaction time test after trying, it was due to difficulty shaking the iPhone hard enough to register a response from the test. Users received an “X” on their screen each time they failed the test, and the research staff identified that the device was being shaken, but “X’s” still appeared due to the low amplitude low force shaking.

#### Completion

Figure 2 presents the results of the multivariable/adjusted and univariate models examining the association between task completion and patient cognition, functional independence, and lack of mobility assistive device usage. Lack of possible cognitive impairment (Six Item Screener score 4-6) was associated with completion rates for the Stroop (*OR* = 4.95, *p* < 0.01) and Trail-Making test times (*OR* = 4.76, *p* < 0.01), but not the reaction time test (Figure 2A). Completion of the reaction time, Gait and Balance, and Timed Walk tests were associated with complete functional independence in univariate but not adjusted models (Figure 2B). Patient cognition, functional independence, and lack of mobility assistive device usage were not associated with completion of the gait and balance test or timed walk test in adjusted models (Figure 2A, B, C).

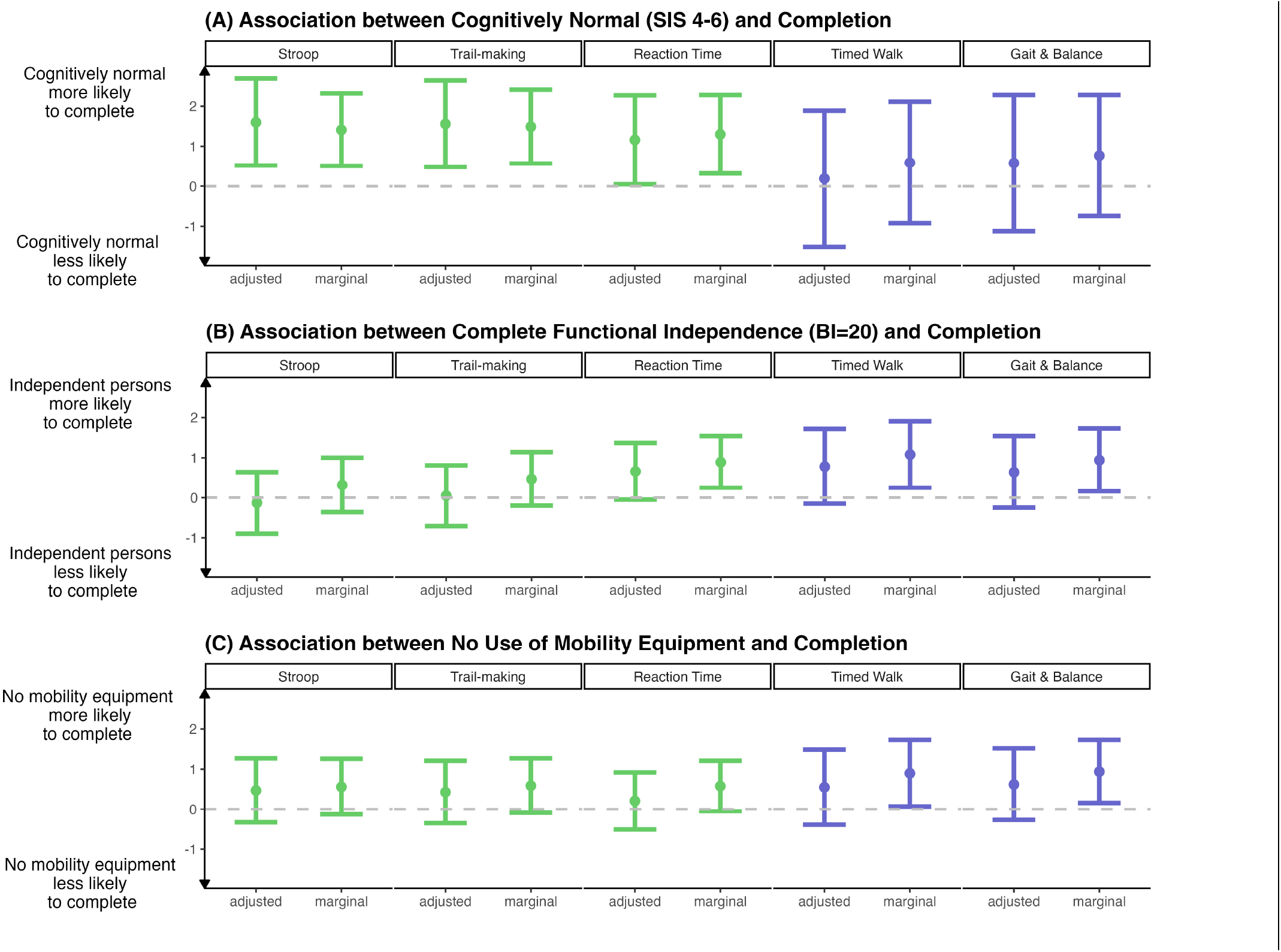

### Univariate Associations of Task Completion with Patient Characteristics

Figure 3 presents a heatmap showing the association of patient characteristics with outcomes in univariate and adjusted models, with outcomes in columns and predictors in rows. Supplementary Tables 1 and 2 provide full details on effect sizes and 95% confidence intervals.

**Figure 3.**
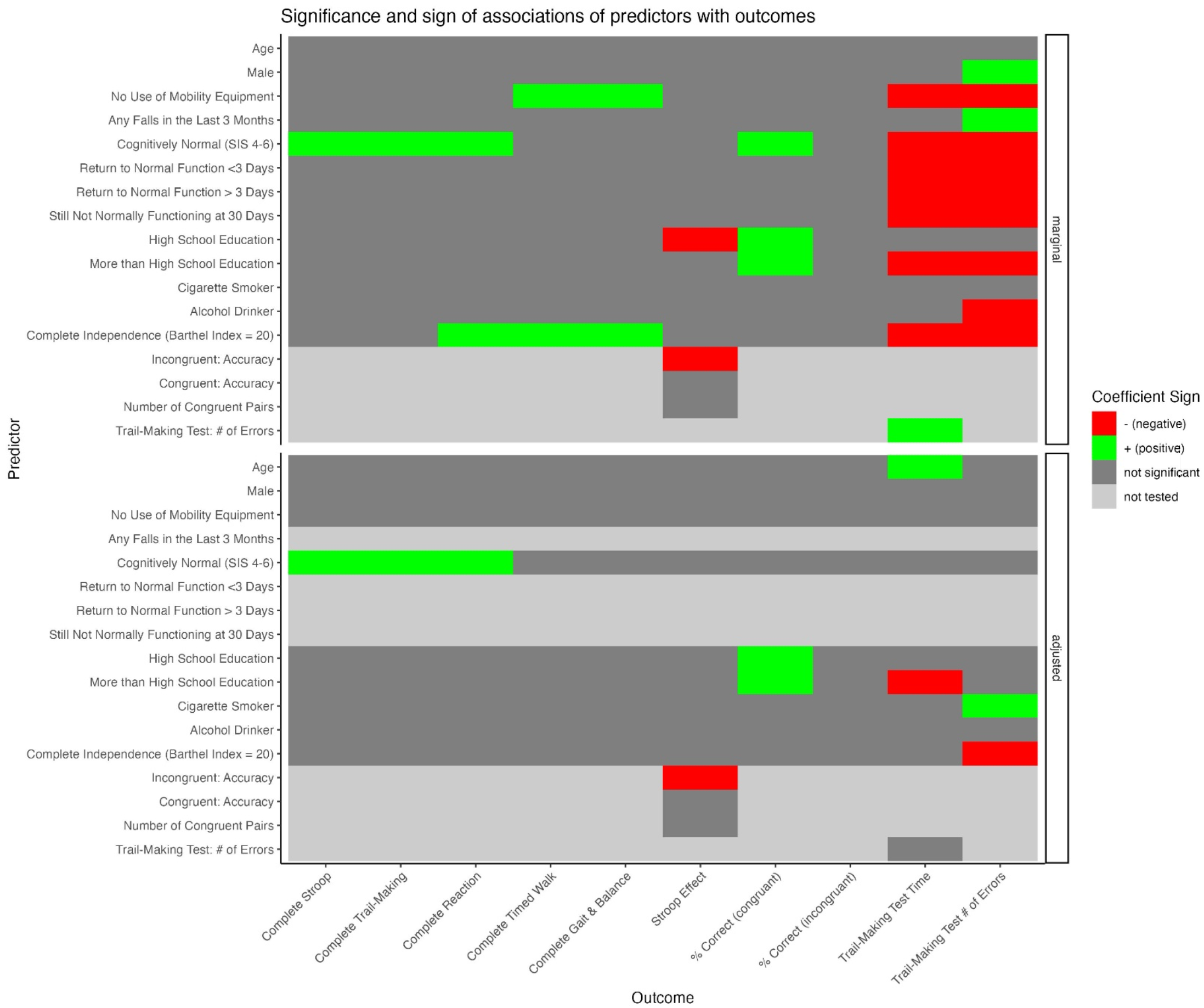
Digital Measures Heatmap showing association of patient characteristics with outcomes in univariate and adjusted models

#### Task Completion

Results for task completion are presented in columns 1-5 of Supplementary Tables 1 (univariate associations) and 2 (multivariable associations). We found that patients with a Six Item Screener of 4-6, indicating probable normal cognition, were more likely to complete the Stroop (OR = 4.10, p<0.01) and Trail-Making tests (OR = 4.44, p<0.01). Cognitive impairment (Six-Item Screener > 4) was similarly associated with completion of the reaction time test (OR = 3.7, p<0.01), as was some level of dependence (modified Barthel Index <20, OR=0.41, p<0.01). Completion of the gait and balance and timed walk tests were similarly associated with functional dependence (OR = 0.39, p=0.02 and OR=0.41, p=0.01, respectively) as well as the use of mobility equipment (OR=0.39, p=0.02 and OR=0.34, p=0.03, respectively).

#### Test Results

Compared to patients with less than a high school education, patients with a high school equivalent education were estimated to have a lower Stroop effect 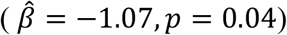 Higher accuracy on the incongruent pairs was also associated with a lower Stroop effect 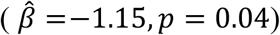. No significant associations were found with accuracy rates for incongruent pairs, though a lack of cognitive impairment (Six Item Screener score 4-6, 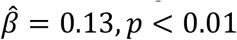 and more than high school education 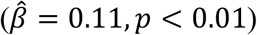 was associated with increased accuracy of congruent pairs for the Stroop test. Increased Trail-Making test time and a larger number of errors on the Trail-Making test were associated with the use of mobility equipment and some functional dependence (modified Barthel Index < 20). No reported recent alcohol consumption, history of falls in the last 3 months, and male gender were associated with a higher number of errors on the Trail-Making test.

### Multivariable/Adjusted Associations of Task Completion with Patient Characteristics

Due to multicollinearity and quasi-complete separation for binary outcomes (completion rates), a reduced set of predictors was considered for multivariable regression models. The predictors included in multivariable regression models were age, gender, use of mobility equipment, cognitive impairment, education, cigarette smoking, alcohol consumption, and modified Barthel Index.

#### Cognitive Test Results

Results for the Stroop effect mirror the univariate results, as high school education 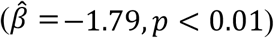, and the number of congruent pairs presented 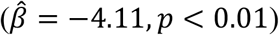 were negatively associated with the estimated Stroop effect. However, lack of cognitive impairment was also positively associated with the Stroop effect, adjusting for other predictors 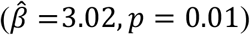. In multivariable regression, lack of possible cognitive impairment was also associated with a decreased accuracy for incongruent pairs 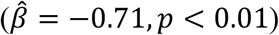 After adjustment for other predictors, only high school education 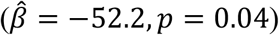 and number of errors 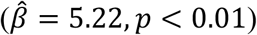 was associated with Trail-Making test completion time. Finally, in adjusted models, lack of cognitive impairment and cigarette smoking were both associated with an increased number of errors on the Trail-Making test

## Discussion

Our study demonstrates that most older adults presenting to the ED after a fall can participate in digital cognitive measures with the assistance of research staff. However, digital assessment of physical function that requires mobilization had fewer completions, primarily due to patients’ reluctance or inability to perform tests immediately after a fall. Our study includes lessons learned for clinicians and investigators who plan to use patient facing digital assessments which can inform the design of future studies and improvement of currently available assessment technologies.

The feasibility of digital cognitive assessments in the ED setting is an encouraging result, indicating that these tools can be integrated into clinical workflows to assess cognitive function in older adults. This finding is consistent with prior research showing high completion of ecological momentary cognitive tests (EMCT) in older adults, with completions ranging from 60-85%, and performance correlated to standard neuropsychological testing scores.^14^ Our study found modestly high completion of the Stroop test and the Trail-Making test, suggesting that similar tools could be effectively utilized in the ED.

However, our findings contrast with prior studies on physical function assessments, which report more completions in non-ED settings. Rubin et al. found that 88% of patients completed a six-minute walk test in perioperative patients using a similar approach,^15^ while our study observed much lower completions of physical function tests (18-20%) immediately post-injury. This difference between our study and Rubin et al.’s likely is due to the timing of tests immediately following an injury and the patients’ conditions as a result of the fall which brought them to the ED. While these low completion rates create missingness and are undesirable, they cannot be solely attributed to the design or administration of the digital tests. Our prior study assessing a traditional test of function, the timed up and go (TUG), also demonstrated low completions among ED patients after falls.^16^ Furthermore, failure to complete the TUG was associated with living in a nursing home six months after the ED visit. Thus, low completion of digital measures requiring mobilization after a fall may be an indicator of fear of falling, frailty, or predictors of future institutionalization rather than an indicator of poor digital measure design or issues with execution.

### Potential implications

Digital cognitive assessments can complement traditional testing, offering a scalable and less resource-intensive option for initial evaluations in the ED. Given the association of cognitive impairment with fall risk,^5^ these tools could help in developing targeted interventions. The findings also underscore the necessity for tailored approaches to physical function assessments, suggesting that patient-specific factors, such as immediate post-fall condition, should be considered when designing such evaluations.

Further, our study provides valuable information to clinicians and investigators who intend to use digital measures in their practice or research. It is valuable to identify reasons for non-attempt and non-completion to distinguish when there was a technology glitch or when completion was not possible due to patient factors. For instance, in our study, 7.2% of measure attempts were incomplete due to technology factors, but the vast majority of noncompletion was because a patient declined testing or initiated testing and then aborted early. Incorporating digital measures into clinical trial design or practice requires the ability to troubleshoot devices, work closely with developers to address changes in app function due to system upgrades or other version issues, and close staff oversight to ensure that recurrent issues are addressed before recruiting new patients. The vigilance required can add complexity to clinical trial delivery and integration of digital tests into clinical practice.

Our trial recruited individuals over four years. Prior to app use and implementation of digital measures, we had to navigate several institutional, regulatory, and technological hurdles. For instance, our data collection software, REDCap, receives data via an application programming interface (API). However, our institution initially did not have REDCap API enabled. This data transfer arrangement required providing evidence to research committees, institutional review boards, and executive sponsors that API could be safely enabled and would be worth enabling due to the increase in sensor use in clinical research that was likely to occur at the institution. We had to identify a developer competent in Xcode, the coding language used to program Apple apps. We also had to do preliminary testing of the ActiveTasks to ensure data generated during our tests was correctly captured and uploaded into the data collection software. Finally, during the four-year study period several iOS updates impacted the ability to open the app and the format of measure data that had to be overcome. These and other challenges were surmountable and are better described in a related manuscript on leveraging ResearchKit for sensor data in clinical trials,^17^ but required dedicated attention aside from the recruitment of older individuals into a multimodal intervention study, adding complexity to the clinical trial oversight.

Eighty nine percent of adults ages 50-64 years and 76% of those over 64 own a smartphone, making digital measures more and more accessible across the lifespan.^18^ A strength of our study was using a single study phone to assess cognition and function and the availability of research staff to provide assistance and training to older adults lacking experience in touch screen use. However, we did note that some older adults lacked the strength to shake the smartphone for the reaction time test vigorously. Thus, noncompletion of this test may be an indicator of frailty or upper extremity strength, in addition to reaction time.

### Why is this important

With the increase in smartphone use in older adults, clinicians and researchers have opportunities to integrate wearable use into practice and research to obtain a better understanding of what occurs to health-related measures after a clinic visit or intervention and in the comfort of patients’ homes. Cognition and physical function are key measures of importance in older adults and our study provides a model for how they can be incorporated into clinical trials, what completion rates can be expected, and how to track noncompletion. Validation studies that show associations between validated “traditional” measures and digital measures are helpful to create uniformity and increase certainty of study findings. Also, digital measures may provide unique and valuable insights and measures into human health. We recommend clinicians and researchers who plan to use wearable devices in their work establish relationships with engineers, computer scientists, developers, and biostatisticians with expertise in analyzing sensor data and develop protocols to ensure uniformity in measure testing, staff procedures, and manuals for patient use of measures. Collecting reasons for nonattempts and noncompletion is valuable to understanding the limitations of the current technology and the human operator. We plan to use this data to refine our study protocol and provide feedback to measure developers to ensure future studies improve on our existing completion rates.

### Future directions

We reported on measures collected during the ED visit only, but our future work will include repeated measures to evaluate intraindividual differences in cognition and function over time and recovery trajectories. Measures obtained in patients’ homes in 12-month follow-up could differ from index measures immediately after the injury, which could reveal important insights on injury recovery and cognitive and functional trajectories after falls. We also plan to compare differences in cognitive and physical function trajectories in the intervention and control group, as patients receiving the GAPcare intervention are known to have fewer subsequent fall-related ED visits ^￼^ and are likely to face fewer declines in their cognition and physical function after the ED visit.

#### Limitations

Although our study is the largest study to date of digital measure assessment in ED patients with falls, several limitations should be noted. Technical issues and patient reluctance contributed to non-attempts and non-completion rates, indicating areas for improvement in both technology and patient engagement strategies. Performing research in the ED during the course of clinical care is challenged by interruptions for clinical priorities (e.g., imaging tests, administration of medication, specialist consultation), and placing additional demands on patients to perform consent, answer surveys, and participate in interventions. We suspect our completion rates may be lower than in future studies, as patients in our clinical trial also underwent consultation by pharmacist and physical therapists, which was time consuming and could have led to fatigue or reduced interest in undergoing more assessments. Although these digital tests mirror many parameters of the gold standard versions of these measures, these digital tests have not been validated against the gold standards and differ from their traditional counterpart. Because of these concerns, it is possible that the digital tests are measuring different or novel constructs or are less sensitive to measuring the same constructs than their more commonly studied and well-validated counterparts. Because the Stroop test was not uniformly applied in terms of the number of congruent/incongruent pairs, chance imbalance could induce spurious associations in the outcomes, which might not be fully accounted for by simple regression adjustment. The external validity of our findings might be constrained by the specific demographic and clinical characteristics of our study population. These findings can be considered a preliminary step towards integrating digital measures in the ED, with further research needed to optimize these tools and assess their long-term impact on patient outcomes.

## Conclusion

Our study provides insight into incorporating digital cognitive and physical function assessments in the ED, highlighting both the potential and the challenges of these technologies. Future research should focus on validating these tools among demographically diverse patient populations, validating digital measures relative to standard (paper) tests, exploring their predictive value for clinical outcomes, and refining methods to enhance completion rates, particularly for physical function assessments. The integration of digital measures in clinical trials could standardize assessments and reduce staff burden, contributing to more efficient and effective patient care.

## Data Availability

Data used in the present study are not currently available for dissemination

---

**Supplementary Table S1:** Univariate regression results. Dependent and independent variables are presented in columns and rows, respectively. Point estimates for regression coefficients (95% CIs) are shown.

|  | Complete Stroop | Complete TMT | Complete Reaction | Complete G/B | Complete Timed Walk | Stroop Effect | Incongruent: Accuracy | Congruent: Accuracy | TMT Time | TMT Errors |
| --- | --- | --- | --- | --- | --- | --- | --- | --- | --- | --- |
| Age | -0.02<br>(-0.06,0.02) | -0.02<br>(-0.06,0.02) | -0.02<br>(-0.06,0.02) | -0.03<br>(-0.08,0.02) | -0.02<br>(-0.07,0.04) | -0.00<br>(-0.04,0.04) | 0.00<br>(-0.00,0.01) | -0.00+<br>(-0.00,0.00) | 2.37+<br>(-0.14,4.88) | 0.00<br>(-0.01,0.01) |
| Male | -0.22<br>(-0.86,0.42) | -0.06<br>(-0.69,0.57) | 0.04<br>(-0.57,0.64) | 0.38<br>(-0.42,1.17) | 0.27<br>(-0.57,1.12) | 0.26<br>(-0.40,0.91) | -0.05<br>(-0.16,0.06) | -0.00<br>(-0.04,0.03) | 17.97<br>(-24.41,60.35) | 0.22**<br>(0.06,0.38) |
| Use Mobility Equipment | -0.56<br>(-1.26,0.13) | -0.59+<br>(-1.27,0.09) | -0.58+<br>(-1.21,0.05) | -0.94*<br>(-1.73,-0.16) | -0.90*<br>(-1.73,-0.07) | -0.29<br>(-0.95,0.36) | 0.04<br>(-0.07,0.15) | 0.01<br>(-0.03,0.05) | 48.70*<br>(6.91,90.50) | 0.44***<br>(0.25,0.62) |
| Any Falls in the last 3 months | -0.31<br>(-0.92,0.30) | -0.32<br>(-0.92,0.28) | -0.37<br>(-0.93,0.20) | -0.05<br>(-0.82,0.72) | -0.02<br>(-0.84,0.79) | 0.29<br>(-0.33,0.91) | -0.07<br>(-0.17,0.03) | -0.00<br>(-0.04,0.03) | 7.91<br>(-32.28,48.11) | 0.36***<br>(0.20,0.52) |
| SIS 4-6 | 1.41**<br>(0.51,2.32) | 1.49**<br>(0.57,2.41) | 1.30**<br>(0.32,2.28) | 0.77<br>(-0.74,2.28) | 0.59<br>(-0.92,2.11) | 0.29<br>(-0.90,1.49) | -0.10<br>(-0.31,0.11) | 0.13***<br>(0.06,0.20) | -160.23***<br>(-236.44,-84.02) | -0.89***<br>(-1.11,-0.66) |
| Return to Normal Function <3 Days | -0.73<br>(-1.98,0.52) | -0.48<br>(-1.63,0.68) | 0.44<br>(-0.67,1.54) | 0.78<br>(-0.96,2.52) | 1.75<br>(-0.45,3.94) | 0.33<br>(-0.90,1.56) | 0.08<br>(-0.14,0.30) | 0.04<br>(-0.03,0.12) | -102.52*<br>(-180.66,-24.38) | -0.72***<br>(-1.02,-0.41) |
| Return to Normal Function > 3 Days | 0.09<br>(-1.21,1.39) | 0.15<br>(-1.02,1.33) | 0.65<br>(-0.44,1.73) | 1.21<br>(-0.44,2.87) | 1.78<br>(-0.39,3.95) | 1.15+<br>(-0.01,2.32) | -0.01<br>(-0.22,0.20) | -0.03<br>(-0.10,0.05) | -118.60**<br>(-194.46,-42.75) | -0.55***<br>(-0.83,-0.27) |
| Still Not Normally Functioning at 30 Days | -0.41<br>(-1.54,0.71) | -0.09<br>(-1.12,0.94) | 0.43<br>(-0.53,1.38) | 0.54<br>(-1.07,2.15) | 1.16<br>(-0.97,3.30) | 0.68<br>(-0.45,1.81) | 0.03<br>(-0.17,0.23) | 0.01<br>(-0.06,0.09) | -91.00*<br>(-163.53,-18.47) | -0.36**<br>(-0.62,-0.11) |
| High School | -0.40<br>(-1.41,0.61) | 0.30<br>(-0.67,1.28) | -0.39<br>(-1.35,0.58) | -0.24<br>(-1.54,1.06) | 0.15<br>(-1.37,1.67) | -1.07*<br>(-2.09,-0.05) | 0.06<br>(-0.12,0.23) | 0.07*<br>(0.02,0.13) | -31.01<br>(-79.70,17.68) | 0.00<br>(-0.26,0.27) |
| More than High School | 0.21<br>(-0.68,1.10) | 0.74+<br>(-0.10,1.58) | 0.13<br>(-0.70,0.95) | -0.07<br>(-1.15,1.01) | 0.47<br>(-0.82,1.76) | -0.03<br>(-0.90,0.83) | 0.03<br>(-0.12,0.18) | 0.11***<br>(0.06,0.16) | -98.07***<br>(-139.05,-57.09) | -0.24*<br>(-0.47,-0.02) |
| Cigarette Smoker | -0.06<br>(-1.78,1.67) | 0.06<br>(-1.67,1.78) | 0.60<br>(-1.13,2.32) | -14.97<br>(-1934.97,1905.03) | -14.81<br>(-1934.81,1905.19) | -1.01<br>(-2.69,0.68) | -0.15<br>(-0.44,0.13) | -0.01<br>(-0.11,0.09) | 37.80<br>(-68.68,144.28) | 0.25<br>(-0.11,0.62) |
| Alcohol Drinker | 0.68<br>(-0.17,1.52) | 0.81+<br>(-0.04,1.65) | 0.53<br>(-0.20,1.26) | 0.82+<br>(-0.04,1.67) | 0.84+<br>(-0.05,1.74) | -0.11<br>(-0.85,0.63) | 0.00<br>(-0.12,0.13) | -0.00<br>(-0.05,0.04) | -7.84<br>(-54.66,38.98) | -0.23*<br>(-0.42,-0.03) |
| Barthel Index < 20 | -0.32 | -0.47 | -0.89** | -0.94*<br>(-1.73,-0.16) | -1.08*<br>(-1.91,-0.25) | -0.15 | 0.02<br>(-0.08,0.13) | -0.01 | 49.94*<br>(9.06,90.82) | 0.54*** |
|  | (-1.00,0.36) | (-1.14,0.20) | (-1.54,-0.25) |  |  | (-0.80,0.51) |  | (-0.05,0.03) |  | (0.35,0.72) |
| Incongruent: Accuracy | - | - | - | - | - | -1.15*<br>(-2.24,-0.06) | - | - | - | - |

\*\*\* p<0.001, \*\* p<0.01, \*p<0.05, +p<0.10

**Supplementary Table S2:** Multivariable regression results. Dependent and independent variables are presented in columns and rows, respectively. Point estimates for regression coefficients (95% CIs) are shown.

| coefficient | Complete Stroop | Complete TMT | Complete Reaction | Complete G/B | Complete Timed Walk | Stroop Effect | Incongruent: Accuracy | Congruent: Accuracy | TMT Time | TMT Errors |
| --- | --- | --- | --- | --- | --- | --- | --- | --- | --- | --- |
| Age | 0.01<br>(-0.03,0.06) | 0.01<br>(-0.04,0.05) | 0.00<br>(-0.04,0.05) | -0.04<br>(-0.10,0.03) | -0.02<br>(-0.09,0.04) | -0.00<br>(-0.04,0.04) | 0.00<br>(-0.01,0.01) | -0.00<br>(-0.00,0.00) | 2.16*<br>(0.31,4.01) | -0.01<br>(-0.02,0.01) |
| Male | -0.41<br>(-1.14,0.32) | -0.20<br>(-0.92,0.52) | 0.05<br>(-0.63,0.73) | 0.40<br>(-0.48,1.28) | 0.25<br>(-0.69,1.18) | -0.07<br>(-0.70,0.55) | -0.08<br>(-0.20,0.04) | -0.01<br>(-0.05,0.03) | 0.98<br>(-29.81,31.76) | 0.03<br>(-0.16,0.23) |
| Use Mobility Equipment | -0.47<br>(-1.27,0.33) | -0.43<br>(-1.21,0.35) | -0.21<br>(-0.92,0.51) | -0.62<br>(-1.52,0.27) | -0.55<br>(-1.49,0.39) | -0.20<br>(-0.87,0.47) | 0.05<br>(-0.07,0.18) | 0.02<br>(-0.02,0.06) | 8.79<br>(-23.95,41.53) | 0.10<br>(-0.11,0.31) |
| SIS 4-6 | 1.60**<br>(0.52,2.69) | 1.56**<br>(0.48,2.64) | 1.16*<br>(0.05,2.27) | 0.58<br>(-1.12,2.28) | 0.19<br>(-1.51,1.89) | 0.14<br>(-1.15,1.42) | -0.12<br>(-0.36,0.12) | 0.07+<br>(-0.01,0.15) | 14.99<br>(-50.69,80.66) | -0.32+<br>(-0.66,0.02) |
| High School | -0.77<br>(-1.88,0.35) | 0.08<br>(-0.97,1.13) | -0.65<br>(-1.68,0.39) | -0.38<br>(-1.76,1.00) | 0.09<br>(-1.50,1.67) | -0.96+<br>(-1.93,0.01) | 0.08<br>(-0.10,0.26) | 0.06*<br>(0.00,0.12) | -39.11<br>(-87.77,9.54) | 0.03<br>(-0.24,0.31) |
| More than High School | -0.37<br>(-1.39,0.65) | 0.24<br>(-0.69,1.17) | -0.40<br>(-1.32,0.52) | -0.60<br>(-1.81,0.61) | 0.07<br>(-1.35,1.48) | -0.20<br>(-1.10,0.70) | 0.07<br>(-0.09,0.23) | 0.09**<br>(0.04,0.15) | -94.42***<br>(-138.40,-50.43) | -0.20<br>(-0.45,0.06) |
| Cigarette Smoker | 0.54<br>(-1.73,2.81) | 0.49<br>(-1.78,2.77) | 1.12<br>(-1.15,3.39) | -15.75<br>(-1894.73,1863.23) | -15.39<br>(-1957.62,1926.84) | -1.16<br>(-2.68,0.36) | -0.14<br>(-0.43,0.15) | -0.03<br>(-0.13,0.06) | 63.61+<br>(-9.03,136.26) | 0.42*<br>(0.04,0.80) |
| Alcohol Drinker | 0.70<br>(-0.24,1.64) | 0.73<br>(-0.21,1.67) | 0.43<br>(-0.39,1.25) | 0.86+<br>(-0.11,1.82) | 0.77<br>(-0.22,1.76) | -0.01<br>(-0.71,0.70) | 0.03<br>(-0.10,0.16) | -0.00<br>(-0.05,0.04) | 16.11<br>(-18.11,50.34) | -0.03<br>(-0.25,0.19) |
| Barthel Index < 20 | 0.13<br>(-0.64,0.90) | -0.05<br>(-0.81,0.71) | -0.66+<br>(-1.37,0.05) | -0.64<br>(-1.54,0.25) | -0.78+<br>(-1.72,0.15) | -0.24<br>(-0.90,0.42) | 0.00<br>(-0.12,0.13) | -0.00<br>(-0.04,0.04) | 27.99+<br>(-3.64,59.62) | 0.39***<br>(0.19,0.59) |
| Incongruent: Accuracy | - | - | - | - | - | -1.74**<br>(-2.78,-0.71) | - | - | - | - |
| Congruent: Accuracy | - | - | - | - | - | 0.20<br>(-2.92,3.32) | - | - | - | - |
| Number of Congruent Pairs | - | - | - | - | - | -1.43<br>(-3.91,1.04) | - | - | - | - |
| TMT Errors | - | - | - | - | - | - | - | - | 2.96+<br>(-0.13,6.06) | - |

\*\*\* p<0.001, \*\* p<0.01, \*p<0.05, +p<0.10

